# Differential inclusion of *NEB* exons 143 and 144 provides insight into *NEB*-related myopathy variant interpretation and disease manifestation

**DOI:** 10.1101/2024.03.25.24304535

**Authors:** Sarah Silverstein, Rotem Orbach, Safoora Syeda, A Reghan Foley, Svetlana Gorokhova, Katherine G. Meilleur, Meganne E. Leach, Prech Uapinyoying, Katherine R Chao, Sandra Donkervoort, Carsten G. Bönnemann

## Abstract

Biallelic pathogenic variants in the gene encoding nebulin (*NEB*) are a known cause of congenital myopathy. We present two individuals with congenital myopathy and compound heterozygous variants (NM_001271208.2: c.2079C>A; p.(Cys693Ter) and c.21522+3A>G) in *NEB.* Transcriptomic sequencing on patient muscle revealed that the extended splice variant c.21522+3A>G causes exon 144 skipping. Nebulin isoforms containing exon 144 are known to be mutually exclusive with isoforms containing exon 143, and these isoforms are differentially expressed during development and in adult skeletal muscles. Patients MRIs were compared to the known pattern of relative abundance of these two isoforms in muscle. We propose that the pattern of muscle involvement in these patients better fits the distribution of exon 144-containing isoforms in muscle than with previously published MRI findings in *NEB*-related disease due to other variants. To our knowledge this is the first report hypothesizing disease pathogenesis through the alteration of isoform distributions in muscle.

## Introduction

Nemaline myopathies are a clinically heterogenous group of skeletal muscle diseases ranging from severe congenital-onset disease to milder childhood-onset disease. Biallelic pathogenic variants in *NEB* (OMIM 161650), a gene encoding nebulin, are a known cause for nemaline myopathy (OMIM 256030), belonging to a histologically defined subgroup of congenital myopathies (Lehtokari et al., 2014). Nebulin is a giant protein (600-900 kDa) that is an important part of the sarcomeric thin filament and is involved in muscle contraction, sarcomere maintenance and homeostasis, although the precise mechanisms of its various functions are still under investigation (Yuen & Ottenheijm, 2020). Nemaline myopathy usually presents congenitally with axial and proximal muscle weakness, prominent facial and bulbar weakness, and respiratory muscle weakness (Lehtokari et al., 2014). Milder phenotypes of nemaline myopathy with onset during childhood or adulthood presenting with distal muscle weakness have also been reported (Lehtokari et al., 2014; Sewry et al., 2019; Wallgren-Pettersson & Laing, 2000). Pathogenic *NEB* variants are a major cause of nemaline myopathy, however, given *NEB*’s 183 exon expanse, and the complexity of its resulting isoforms, the identification of variants is challenging, and the functional impact of *NEB* variants is difficult to interpret.

Alternative splicing creates diversity of function from an otherwise limited number of genes. Differential splicing programs are well-established within skeletal muscle for various developmental states and between type I and type II myofibers; perturbations to these alternative isoform programs are known to drive muscle disease (Nakka et al., 2018, Trapnell et al., 2010, Matyushenko et al., 2020; Nikonova et al., 2020; Robaszkiewicz et al., 2020). Moreover, alternative splicing characteristics between mature muscles that are not associated with fiber type composition or development have recently been explored for titin encoding gene (*TTN*), although disruptions in the relative abundance of these isoforms have not yet been linked to disease (Freiburg et al., 2000; Nikonova et al., 2020; Savarese et al., 2018). .

*NEB (*NM_001271208.2) is a complex transcriptional unit and is known to include alternatively spliced exons (143 and 144) that are mutually exclusive. Exon 143 is thought to be prominently expressed in the fetal period, while exon 144 is shown to be expressed with increasing age in both mice and humans (Donner et al., 2004, 2006; Laitila et al., 2012; Lam et al., 2018; Uapinyoying et al., 2020). While certain adult muscles ultimately express an equal ratio of both exons (e.g., vastus intermedius and medialis) others express higher amounts of exon 143 compared to exon 144 (e.g. tibialis anterior), and some adult muscles exclusively express exon 144 (e.g. gastrocnemius, rectus femoris) (Laitila et al., 2012). As such, a role for the specific isoforms in myogenesis and fiber typing is postulated (Lehtokari et al., 2014), however the impact on disease mechanism and disease manifestation in humans has not yet been explored.

RNA sequencing has emerged as a promising tool for understanding the impact of specific variants on transcription and splice regulation, thus aiding in assessing pathogenicity. Here we report detailed phenotypic data on two siblings affected with congenital onset muscle weakness in whom we identified compound heterozygous pathogenic variants in *NEB (*NM_001271208.2: c.2079C>A; p.(Cys693Ter) and c.21522+3A>G). Aberrant splicing of the *NEB* intronic variant was confirmed by muscle RNA sequencing. We also explore how the differential expression of *NEB* isoforms may be linked to muscle function in normal and diseased muscle. Further, we propose that differential isoform expression across muscle groups may explain disease-specific patterns of muscle involvement in skeletal muscle disease.

## Subjects and Methods

### Patient recruitment and sample collection

Patients were referred by their neurologist or geneticist. Written informed consent and age-appropriate assent for study procedures were obtained by a qualified investigator (protocol 12-N-0095 approved by the National Institute of Neurological Disorders and Stroke, National Institutes of Health Institutional Review Board (IRB)). Medical history was obtained and clinical evaluations, including muscle magnetic resonance imaging (MRI) and muscle biopsy, were performed as part of the standard diagnostic evaluation. Blood samples for research-based testing were obtained using standard procedures.

### Genetic testing

Quartet exome sequencing was performed on DNA isolated from whole blood. Exome sequencing and data processing were performed by the Genomics Platform at the Broad Institute of MIT and Harvard with an Illumina exome capture (38 Mb target) and sequenced (150 bp paired reads) to cover >90% of targets at 20x and a mean target coverage of >100x. Exome sequencing data was processed through a pipeline based on Picard and mapping done using the BWA aligner to the human genome build 38. Variants were called using Genome Analysis Toolkit (GATK, HaplotypeCaller package version 3.4).

Human whole transcriptome sequencing was performed by the GenomicsPlatform at the Broad Institute of MIT and Harvard on RNA isolated from patient 1 muscle (biopsy taken in early childhood from biceps muscle). The transcriptome product combines poly(A)-selection of mRNA transcripts with a strand-specific cDNA library preparation, with a mean insert size of 550 bp. Libraries were sequenced on the HiSeq 2500 platform to a minimum depth of 50 million reads. ERCC RNA controls were included for all samples, allowing additional control of variability between samples. Raw sequencing data was aligned using the GTEX pipeline version 10 (https://github.com/broadinstitute/gtex-pipeline/tree/master/rnaseq).

Confirmation of variants discovered in patients and segregation testing in the parents and unaffected sibling was performed by Sanger sequencing.

### Splice Variant Analysis

Patient 1 BAM file was analyzed using the Integrative Genomics Viewer (IGV) (Thorvaldsdottir et al., 2013). Considering the known variability of exon 143 versus 144 inclusions depending on age and muscle biopsy site, we carefully searched for appropriate controls for Patient 1. Unfortunately, controls matching both age and biopsy site were unavailable. We could not use the GTEx muscle samples as controls, as the youngest sample in GTEx is an 18-year-old (yo) individual (with most of the samples 65+), and the muscle samples were from the gastrocnemius muscle or designated as from “below the patella.” Thus, controls were selected from our own disease cohort excluding any sample with known *NEB*-related disease. The only available RNA-seq samples from biceps tissue were from adults. All RNA-seq samples in our cohort with biopsies taken at early childhood are from the quadriceps muscle. We therefore selected two age-matched control samples and two controls matching the biopsy site. Sashimi plots were generated with ggsashimi (Garrido-Martín et al., 2018).

### Transcript Usage

The MANE select and MANE plus Clinical transcripts for *NEB* are NM_001164508.2 and NM_001164507.2, respectively. Although the MANE select transcript is used to report *NEB* variants in ClinVar and gnomAD, we utilized the alternative transcript NM_001271208.2 in this study. The MANE select and MANE plus Clinical transcripts each contains only one of the two mutually exclusive exons 143 and 144 and are thus both labeled as “exon 143” in their respective transcripts. To preserve clarity as we discuss both exons, we use the non-biological NM_001271208.2 transcript, which contains both exons 143 and 144.

### Muscle MRI imaging and relative exon expression analysis

Muscle MRI was performed using conventional T1-weighted spin echo and short tau inversion recovery (STIR) of the lower extremities. Multiple sequences including axial STIR and T1 images were obtained at 3 Tesla without vascular contrast administration.

T1 axial images of proximal and distal lower extremities were reviewed in detail to note pattern of muscle involvement and fibroadipose transformation (Lamminen, 2014; Ortolan et al., 2015) to include a qualitative Modified Mercuri Scoring (MMS) of the thigh and lower legs as previously done by Perry and his colleagues (MMS: 0= Normal appearance; 1= Early fat infiltration, scattered areas of T1 high signal; 2= Numerous discrete areas of T1 high signal with beginning confluence <30% of the volume; 3= Fat infiltration 30–60% of volume; 4= Fat infiltration >60% of the volume; 5= End stage, no residual muscle tissue (Perry et al., 2023). P1 MMS and P2 MMS (calculated on his muscle MRI from teenage years) were averaged per muscle to reflect detailed exon 144 skipping genotype pattern. MRI of age-matched Control *NEB* patients (ie *NEB* patients with variants other than *NEB* c.21522+3A>G) were selected from *NEB* patients reported by Perry et al 2023, and mean MMS was calculated from reported scores (supplemental table 3) (Perry et al., 2023).

Current literature was reviewed to identify the reported relative expression of exon 143 and 144 across lower limb muscles in adult humans and mice (Supplemental Table 2). The collected data was subsequently binned into four qualitative categories per muscle: 1) Exon 144 only, 2) Exon 144 was expressed more than Exon 143, 3) Exon 143 was expressed equally with Exon 144 and 4) majority Exon 143. Where human data conflicted with mouse data, human data was used to make Figure D6. If no human data was available and a conflict found between mouse datasets, the more quantitative dataset from Uapinyoying et al 2020 was used to make Figure D6. The muscle MRI grading for each muscle was then compared to the relative expression of exons 143 and 144.

## Results

### Clinical Presentation

Patient (P1) is a young adult male of American and Asian ancestry. He was born to non-consanguineous parents at 42 weeks gestational age via Cesarian section. Pregnancy was complicated by decreased fetal movements. Early gross motor milestones were attained on time, achieving independent ambulation at the upper limit of normal. Early walking gait was described as wide-based, waddling and with inward rotation of the knees. During childhood he was noted to have facial weakness, while fine motor, language and cognitive development were normal. On physical exam at age in early childhood, he had a high-arched, narrow palate, facial weakness, atrophic gluteal muscles, proximal more than distal muscle weakness and a positive Gowers’ maneuver. Gait was described as wide-based and waddling with exaggerated effort to perform knee extension. Additionally, he was noted to have hyperlaxity in joints of the hands and contractures of the Achilles tendons. Creatine Kinase (CK) levels were reportedly normal throughout childhood. Electromyography studies were consistent with non-irritable myopathy. A muscle biopsy performed in early childhood showed non-specific myopathic changes without nemaline rods on histology (Figure A). Electron microscopy was not available. Echocardiogram and pulmonary function studies were within normal limits. Serial examinations in late childhood and teenage years demonstrated slightly improved muscle strength throughout (Table 1, available on request). At examination as a young adult, the patient reports worsening muscle weakness and stamina along with weight gain, difficulty chewing, frequent falls, myopia, hypertension, and borderline conductive hearing loss secondary to multiple episodes of otitis media. The hypertension and worsening muscle weakness may be the result of significant weight gain.

**Figure A:**
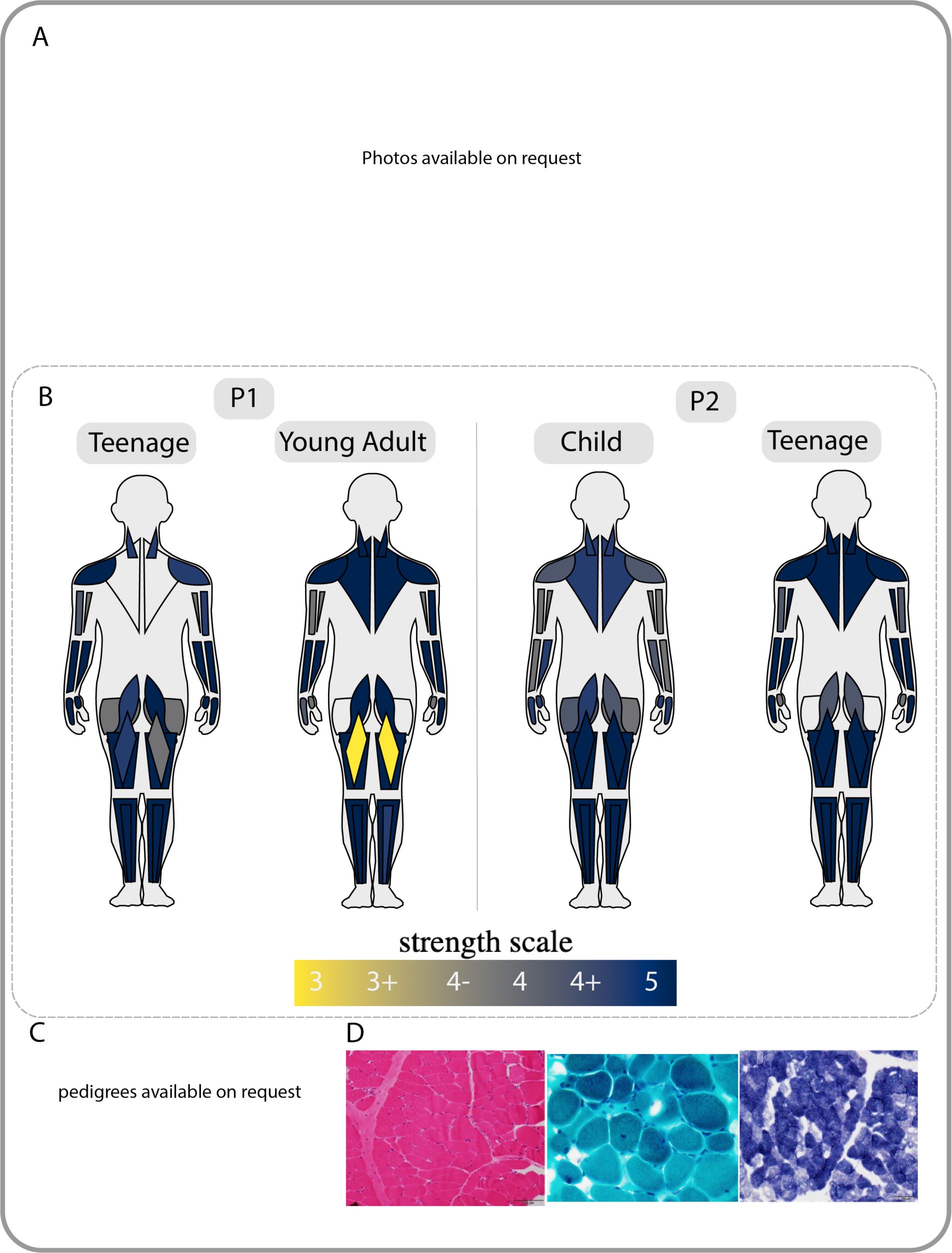
Patient Phenotype and Pedigree A) facial weakness, ankle contractures, narrow palate, and body habitus in P1 (left) and P2 (right). B) strength grading for both P1 and P2 at two timepoints, diagram created by MuscleViz (https://muscleviz.github.io/). C) pedigree and segregation of NEB variants in our patient family. D) histology from P1, H&E (left) Gomori Trichrome (middle) and NADH (right), no nemaline rods noted. EM images were unavailable for analysis.

Patient (P2) is a teenage male. Pregnancy was complicated by decreased fetal movement in one of the twins. He was born at 38 weeks gestational age via C-section due to transverse positioning. There were no concerns at birth, and early milestones were normal. At early childhood, P2’s parents noted that he had difficulty climbing stairs. On physical examination in childhood, he had a transverse smile, atrophic gluteal muscles, proximal muscle weakness and an exaggerated Trendelenburg gait with arm pumping. He had a negative Gowers’ maneuver (Figure A). CK levels were normal. Examination at age late childhood showed slightly improved muscle strength. Electromyography studies performed revealed findings suggestive of a non-irritable myopathy. As a teenager, P2 reported a regular regimen of weight training and stable to improved strength, although noted recent difficulty with chewing tough foods. Echocardiogram and pulmonary function testing have been normal (Table 1, available on request).

### Exome Sequencing identified biallelic variants in NEB

Quartet exome sequencing (ES) was performed, which identified compound heterozygous variants in NEB (NM_001271208.2) in both individuals. The maternally inherited variant (c.2079C>A; p.Cys693Ter) results in an early stop codon in exon 22 and is predicted to induce nonsense mediated decay. This variant is reported in 3 individuals in gnomAD exomes v4.0.0 with a Grpmax filtering allele frequency (European, non-Finnish) of 0.000002280% (Karczewski et al., 2022; Whiffin et al., 2017). The paternally inherited variant (c.21522+3A>G; rs148950085) is adjacent to exon 144. The c.21522+3A>G variant has conflicting interpretations of pathogenicity in ClinVar based on 6 submissions, 4 pathogenic and two Uncertain Significance (Variant ID 430110) and is predicted to result in donor loss via in-silico tools (CADD prediction v1.6 24 (Rentzsch et al., 2021); spliceAI 0.71, (Jaganathan et al., 2019)). This variant has also been reported as pathogenic in several recently published cohorts when in trans with a missense or truncating *NEB* variant, but never in homozygosity (Supplemental Table 1, available on request) (Lee et al., 2017; Wang et al., 2020; Wen et al., 2020). This variant has a Grpmax filtering allele frequency (East Asian) of 0.001567% in gnomAD exomes and genomes v4.0.0 and one homozygote is reported. The presence of one homozygote in gnomAD does not exclude pathogenicity for two reasons. Firstly, the age of this individual is unknown and biallelic *NEB* variants have been reported to cause disease late in life, and secondly, hypomorphic splice variants are reported to cause disease only in trans with a pathogenic variant but not when found in a homozygous state (Zernant et al., 2017). P1 and P2 report paternal Asian ancestry, and it has been suggested that c.21522+3A>G originates from a common Asian ancestor (Lee et al., 2017; Wang et al., 2020; Wen et al., 2020). While the highest population frequency for this variant is found in East Asians, it is also reported in the gnomAD exomes and genomes v4.0.0 database in the South Asian population with a frequency of 0.0002624 (22/ 22766 alleles) to the exclusion of any additional populations (Karczewski et al., 2022; Whiffin et al., 2017). In summary, this suggests that P1 and P2 share compound heterozygous variants which cause nemaline myopathy, one variant which is only reported in East and South Asian populations and the mechanism for disease remains unclear. No other candidate variants were identified by exome sequencing.

### RNA sequencing confirms NEB c.21522+3A>G causes impaired splicing

To evaluate the effect of the biallelic *NEB* variants identified in ES, whole transcriptomic sequencing on P1 muscle was performed. At the location of the c.2079C>A; p.Cys693Ter variant, DNA sequencing shows a balanced (50:50) heterozygous C>A variant while the RNA sequencing shows allelic imbalance in favor of the wild-type allele (83% G to 17% T, Supplemental Figure 1), suggestive of nonsense mediated decay of the maternal allele transcript with the c.2079C>A; p.Cys693Ter variant. Additionally, exon coverage throughout the *NEB* gene seems to be reduced compared to control RNA-seq samples, although this cannot be assessed with certainty without quantification and normalization for library size. At the location of the intronic c.21522+3A>G variant, almost complete exclusion of exon 144 is seen in the patient sample compared to age-matched controls (Figure B and Supplemental Figure 3).

**Figure B:**
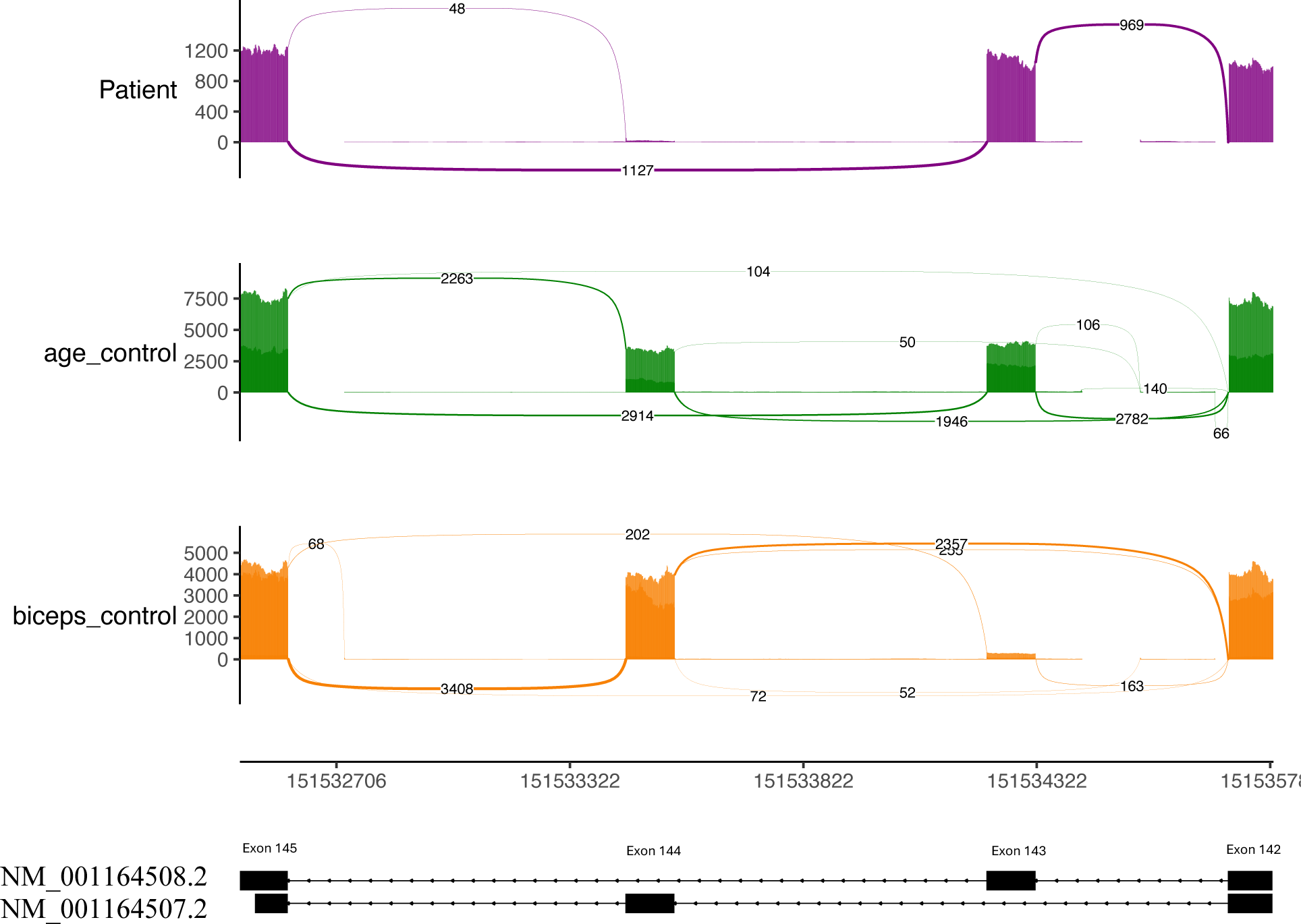
RNA sequencing of NEB exons 143 and 144 in patient P1 and controls. The age control group is the aggregate of two age matched control samples from quadriceps. Reported as median junction reads. The biceps control group is an aggregate of three adult patients with biceps biopsies, aggregate by median junction. The patient biopsy has very few junction reads to exon 144 compared with both controls. Junctions with less than 10 reads were removed for clarity. All junctions and individual control samples can be viewed in supplemental figures 2 and 3.

Although no age- and biopsy-matched samples are available for comparison, RNA-seq data from adult biceps similarly shows that exon 144 is excluded in P1 compared with controls (Supplemental Figure 2). As no relative expression data is reported for biceps (Donner et al., 2004, 2006; Laitila et al., 2012; Lam et al., 2018; Uapinyoying et al., 2020), it is possible that in biceps exon 144 is not included normally. In addition, a developmental difference cannot be excluded as our patient was early childhood at the time of biopsy, whereas the controls were adults. However, there is one report of an 18-year-old female patient with a different base pair change (A>C instead of A>G) at the same site as our variant, causing cryptic donor activation leading to exon 144 extension (Cummings et al., 2017). This was assessed in RNA-seq data obtained from either vastus lateralis or biceps muscle (Cummings et al., 2017). Notably, we observe six novel isoform reads containing exon 144 extension due to the same cryptic donor activation as reported in the A>C variant in our patient sample but not in controls (Supplemental Figure 2 and 3). While the region between exons 142-145 contain multiple splice junctions, the exon 144 exclusion and six reads supporting cryptic donor activation are the only differences in the patient sample compared with controls (Supplemental Figure 2 and 3). Irrespective if the c.21522+3A>G variant causes intron retention or cryptic donor activation with exon extension, we expect that the variant in P1 causes complete loss of the exon 144 containing protein isoform due to a PTC after 7 amino acids, leaving only the exon 143 isoform available for translation (Figure C).

**Figure C:**
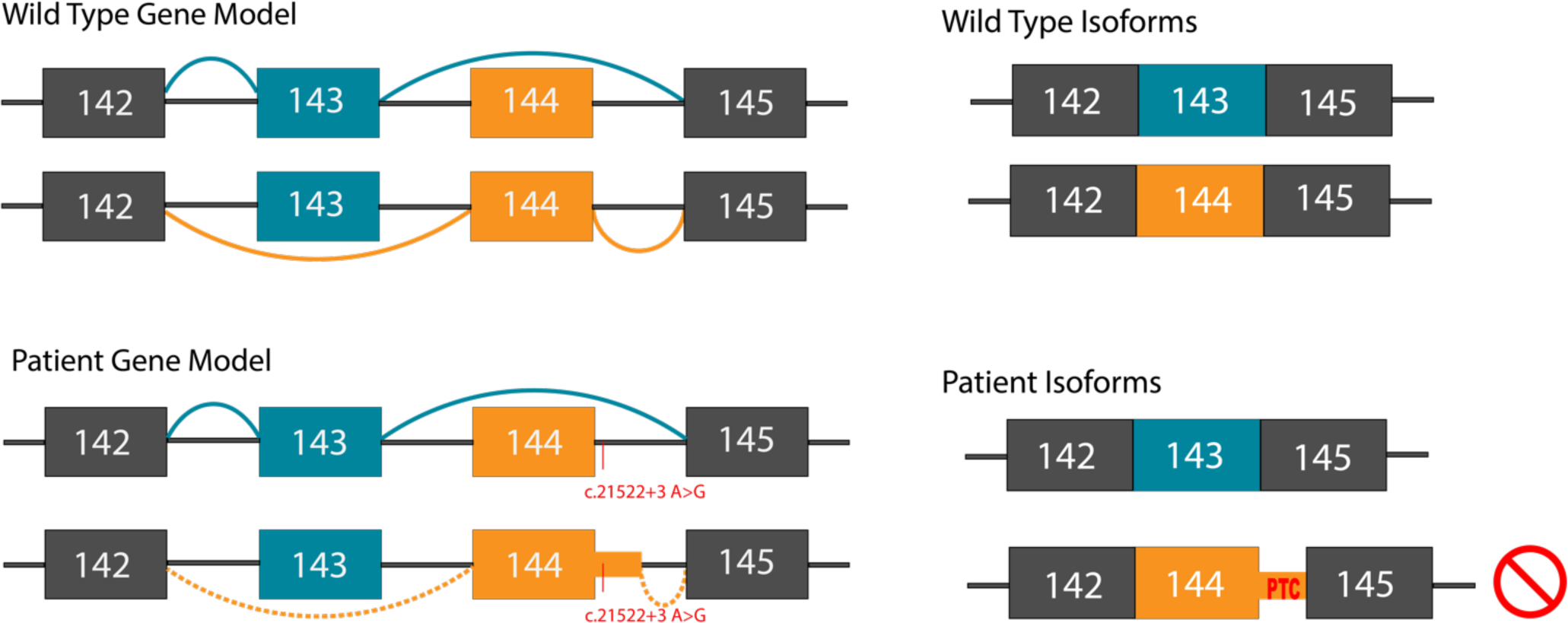
NEB (NM_001271208.2) gene models illustrating the different splicing patterns involving the mutually exclusive exons 143 and 144 in control individuals and patients. The patient variant affects splicing leading to generation of an aberrant exon 144 containing isoform with a premature stop codon resulting in nonsense mediated decay.

### Muscle MRI imaging have a distinct pattern of involvement

Muscle MRI of P1 (as a teenager) and P2 (as a teenager) reveals striking proximal involvement of the vasti muscles, adductor magnus, biceps femoris and semimembranosus (mMMS: 2.5-5) (Figure D, Supplemental Table 3). In contrast, age-matched control NEB patients are relatively spared for the same muscle groups (mMMS: 0.75-1.0) (Perry et al., 2023).

**Figure D:**
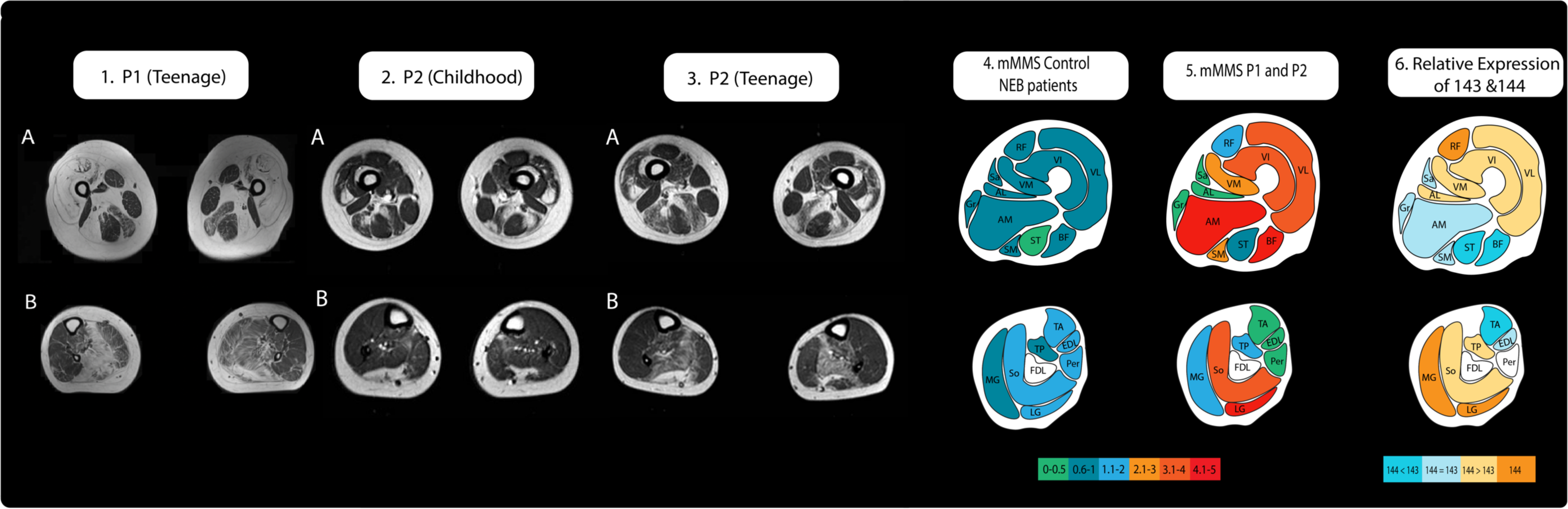
Muscle MRI T1 and relative expression of exons 143 and 144. 1A) Significantly increased T1 signaling, indicating severe, almost complete, replacement of muscle with fibroadipose tissue in the vasti, rectus femoris (RF), long head of biceps femoris (BF), adductors (Ad) with relative sparing of semitendinosus (ST), Sartorius (Sr) and Gracilis (Gr) and milder involvement of the semimembranosus (SM). 1B) In the lower leg there is fibroadipose replacement of lateral gastrocnemius (LG) and soleus (So) (right>left) with relative sparing of anterior compartment muscles. 2A) Increased T1 signaling consistent with fibroadipose replacement in a patchy distribution in the vastus lateralis (VL), vastus intermedius (VI), semimembranosus (SM) and long head of biceps femoris (BF) muscles (left>right) in the upper leg 2B) and patchy fibroadipose replacement of the lateral gastrocnemius (LG) and soleus (So) muscles in the lower leg. 3A) Symmetric patchy fibroadipose replacement of VL and VI with relative sparing of vastus medialis (VM). In the posterior thigh, there is selective and more prominent involvement of SM and long head of BF, slightly more significant on the left compared to the right. RF, Sr, Gr, short head of BF, and ST are selectively spared and appear hypertrophied. 3B)DFibroadipose replacement of LG, So, and tibialis posterior (TP) with sparing of the anterior lower leg muscles and medial gastrocnemius (MG)._J 4) mean Modified Mercuri Scores reported for age matched *NEB* patients (supplemental table 3) (Perry et al, 2023) 5) mean Modified Mercuri Scores reported for P1 and P2 (supplemental table 3) 6) Approximate depiction of relative expression of exons 143 and 144 as reported in the literature_J(Donner et al., 2004, 2006; Laitila et al., 2012; Lam et al., 2018; Uapinyoying et al., 2020).

The rectus femoris, semitendinosus, sartorius, gracilis and adductor longus are relatively spared for both our patients and controls (mMMS P1/P2: 0-2; mMMS control: 0.5-0.75) (Figure D, Supplemental Table 3) (Perry et al., 2023). However, it is worth noting that P1 rectus femoris muscle was extensively replaced by fibroadipose tissue and rated as MMS 4, while P2 rectus femoris muscle was spared and rated as 0 (Supplemental Table 3).

Distally, P1 and P2 have striking involvement of the soleus and lateral gastrocnemius muscles (mMMS: 4-4.25), with sparing of the anterior compartment (mMMS: 0).

Comparatively, control NEB patients presented with mild involvement of the soleus and lateral gastrocnemius (mMMS: 1.25-2) and the anterior compartment (mMMS: 1.5) (Perry et al., 2023). The medial gastrocnemius and tibialis posterior show mild involvement for both our patients and control(s?) (mMMS P1/P2: 1.5-2.0; mMMS control: 0.75-1.0) (Figure D, supplemental table 3) (Perry et al., 2023).

## Discussion

Rare congenital myopathies and dystrophies frequently pose diagnostic challenges considering that several important disease-causing genes expressed in skeletal muscle are exceedingly long and have complex splicing patterns. Here we present two affected individuals each carrying a c.21522+3A>G variant near the mutually exclusive exon 144 of *NEB* (NM_001271208.2) in trans with a truncating NEB variant. A literature review on mutually exclusive exons 143 and 144 suggests that once adulthood is reached, the relative abundance of these two isoforms vary amongst muscles. Many of these muscles exhibit a shift from predominant exon 143 expression early in development to exon 144 with age, although most muscles express a mixture of both isoforms (Figure D6, Supplemental Table 2) (Donner et al., 2004, 2006; Laitila et al., 2012; Lam et al., 2018; Uapinyoying et al., 2020). However, relative abundance was analyzed mostly via qPCR data in mouse tissue, with only a small amount of human muscle contributing, and excepting one study provided no correlation between mRNA abundance and protein levels (Donner et al., 2004, 2006; Laitila et al., 2012; Lam et al., 2018; Uapinyoying et al., 2020). More work to systematically establish the precise distributions in human muscle and elucidate the functional roles needs to be done to aid interpretation of variant pathogenicity in these mutually exclusive exons.

The c.21522+3A>G splice variant found in our patients has recently been reported in patients with nemaline myopathy (Lee et al., 2017; Wang et al., 2020; Wen et al., 2020). Previously, RT-PCR performed on patient muscle cDNA to interpret the functional impact of the c.21522+3A>G variant suggested the generation of a splice isoform that excluded both exons 143 and 144, splicing exon 142 directly to exon 145 (Wang et al., 2020). With RNA sequencing, however, a more complex story emerges as both patient and controls show a few reads splicing exons 142 to 145, suggesting that this is a rare but normal isoform (Supplemental Figure 2 and 3). This is corroborated in GTEx muscle samples (Supplemental Figure 4). Instead, we observe loss of exon 144 and 6 unique reads mapping to a cryptic donor site, suggestive of exon 144 extension leading to nonsense mediated decay (Figure B and supplemental figures 2 and 3).

While we cannot be certain of the precise splice isoform generated due to low read depth at the cryptic donor site in our patient sample, we can be confident that the variant causes loss of exon 144 isoforms when in trans with a nonsense variant (Figure C: Splicing Diagram). Utilizing transcriptomic sequencing for analysis of this complex splice region provided insight into pathogenesis unobtainable through methods like RT-PCR.

Considering the developmental roles of exons 143 and 144 reported in the literature, transcriptomic sequencing can be difficult to interpret in this complex region of *NEB*, and choice of control is especially important (Donner et al., 2004, 2006; Lam et al., 2018). This is illustrated by a variant in a 3-year-old male with congenital myopathy at the +4 site published by Cummings et al., 2017, where RNA sequencing derived from biceps muscle demonstrated switching to exon 143-containing transcripts. However, as one control sample similarly showed this isoform switching, the authors concluded that this is not a pathogenic variant. Examination of supplemental data reveals that this control sample is from a 1-year-old female with unknown biopsy origin and may thus not be sufficient evidence to exclude pathogenicity of the extended splice variant (Cummings et al., 2017). Although the unbiased approach of RNA sequencing can better detect complex splice events, these alternatively spliced isoforms prove difficult to assess considering the variability by age and muscle type in healthy tissue.

Our patients presented with a pattern of muscle involvement atypical to the primarily distal involvement reported for *NEB* nemaline myopathy (Figure D) (Jungbluth et al., 2004; Perry et al., 2023; Scoto et al., 2013). Notably, there is one report of a 54-year-old woman with biallelic variants in *NEB* and primarily proximal muscle involvement (Wunderlich et al., 2018). Since exon 144-containing isoforms are lost in our patients, we thought to examine if the MRI pattern of involvement follow the distribution of exon 144. When comparing our patient muscle MRI to the established literature on the adult relative distribution of exons 143/144 isoforms (Figure D6, Supplemental Table 2, Supplemental Table 3), muscles that are known to contain higher or equal exon 143 expression (sartorius, gracilis, semitendinosus, tibialis anterior and extensor digitorum longus) were shown to be relatively spared while muscles with higher amounts of exon 144 (vastus lateralis, vastus medialis, lateral gastrocnemius and soleus) were more affected (Figure D). Notably, the rectus femoris, biceps femoris, adductors longus/magnus and medial gastrocnemius did not fit with the distribution of exon 144 (Figure D). Intriguingly, the rectus femoris was significantly involved in P1 (MMS=4) while uninvolved in P2 (MMS=0), suggesting that exon 144 expression cannot completely explain P1’s MRI (Supplemental Table 3). Additionally, the long head of the biceps femoris was strikingly involved in our patients (mMMS=4.5), while the short head was uninvolved (mMMS=0), but no data was available regarding which part of the biceps femoris was surveyed in relative abundance studies (Figure D, Supplemental table 3). The adductor magnus is described as containing both isoforms in the literature, with no specific relative abundance for comparison (Figure D6, Supplemental Table 3). The semimembranosus is described as having less 144 than the adductor longus, but no relative expression data is available and thus may well have more 144 than 143, which would fit with the involvement seen in our patients. The overall impression is that the patient MRI findings fit with the known pattern of exon 144 distribution more than with what had been published in the literature. However, one recent published report of childhood onset nemaline myopathy due to a nonsense variant in trans with the c.21522+3A>G variant provides MRI imaging taken in adulthood refuting our analysis (Wen et al., 2020). This patient is reported to have muscle involvement typical of nemaline myopathy, including involvement in the tibialis anterior, although muscle grading is not provided for comparison and the images are insufficient (Wen et al., 2020). Although a functional improvement in strength was noted for P2, the MRI data showed mild progression of fibroadipose replacement of affected muscles, which may be due to the increased reliance these muscles have on exon 144 as adulthood is reached (Figure D2-3). Additional MRI imaging taken at several timepoints from patients with variants impacting exon 144 *NEB* splicing are needed to establish whether perhaps there may be a variant-specific pattern of muscle involvement on MRI.

Understanding the unique roles these mutually exclusive exons play in the sarcomere may be important both to understand muscle development and to identify therapeutic approaches in *NEB*-related myopathy. Considering that the *NEB* gene contains other notable alternatively spliced regions (exons 63-66, 82-105, and 166-177), insight into the function of exons 143 and 144 may inform investigations into these other regions (Donner et al., 2004). Besides their different relative abundance, the mutually exclusive isoformsthat contain exon 143 or 144 differ in charge and hydrophobicity, and exon 144 contains a predicted protein kinase C phosphorylation site (Donner et al., 2004). A step towards gaining insight into the roles of these isoforms may be through uncovering a genotype-phenotype correlation. With the heterogeneity of disease clinical presentation, no genotype-phenotype relationship is currently established for pathogenic variants in *NEB* (Amburgey et al., 2021; Sewry et al., 2019). To date, patients described with the c.21522+3A>G splice variant have been categorized with congenital-onset or mild-onset disease, but none with severe disease (Supplemental Table 1, available on request) (Lee et al., 2017; Wang et al., 2020). Furthermore, a patient with a homozygous deep intronic variant in intron 144 leading to complete loss of exon 144 was reported with a phenotype of congenital-onset nemaline myopathy and proximal more than distal weakness (Laflamme et al., 2021). Given the adult role of exon 144, it is possible that untampered expression of developmentally necessary exon 143 helps maintain the sarcomere and prevents severe disease. Although a consensus exists for the diametric roles of these isoforms (supplemental Table 2), one study challenges this binary understanding (Donner et al., 2006; Lam et al., 2018). When probed in cell culture as opposed to muscle tissue, exon 144 is expressed exclusively in myoblasts, switching to exon 143 expression upon differentiation into early myotubes (Lam et al., 2018). Further work to understand the full pattern of expression will be important to properly understand the function of these exons.

Finally, *NEB-*related myopathy is typically a form of nemaline myopathy (Table 1, available on request), so the absence of the hallmark finding of nemaline rods in the muscle biopsy in our patients is noteworthy (Figure A), since all reported patients with the c.21522+3A>G variant were found to have nemaline rods on biopsy (Lee et al., 2017; Wang et al., 2020; Wen et al., 2020). In nemaline myopathies in general, the presence and percentage of rods in a patient biopsy can be variable and, although diagnostically helpful, may not correlate with disease severity (Sewry et al., 2019; Wang et al., 2020). Furthermore, given the differential muscle involvement evident on imaging, it is conceivable that nemaline rods could indeed be found in another muscle. Thus, the absence of nemaline rod pathology in our patients does not rule out the diagnosis, but highlights the importance of still considering *NEB-*related myopathy in the compatible clinical context, while taking into consideration the many complexities of the *NEB* gene as evident on genomic and transcriptional analysis (Wallgren-Pettersson et al., 2007).

### Future Directions

Additional patients and research are needed, especially to assess how lack of exon 144-containing transcripts affects muscle function and patterns of involvement. Further work to validate the relative abundance of these isoforms in humans and their functional or regulatory impact will better aid our understanding of muscle development and disease pathogenesis.

Finally, while to our knowledge this is the first report of disease pathogenesis through the alteration of isoform distribution, we speculate that more examples of splice active variants skewing alternative splicing may be found, establishing this as a pathogenic mechanism for neuromuscular disease.

## Supplemental Information

Supplemental Information includes three tables and four figures

## Data Availability

All data produced in the present study are available upon reasonable request to the authors

## Acknowledgement

We would like to thank the families for their participation. The work in C.G. Bönnemann’s laboratory is supported by intramural funds from the NIHLJNational Institute of Neurological Disorders. Sequencing and analysis were provided by the Broad Institute of MIT and Harvard Center for Mendelian Genomics (Broad CMG) and was funded by the National Human Genome Research Institute, the National Eye Institute, the National Heart, Lung and Blood Institute and grant UM1HG008900 to Daniel MacArthur and Heidi Rehm. This study makes use of data shared through the seqr platform with funding provided by National Institutes of Health grants R01HG009141 and UM1HG008900.

The Genotype-Tissue Expression (GTEx) Project was supported by the Common Fund of the Office of the Director of the National Institutes of Health, and by NCI, NHGRI, NHLBI, NIDA, NIMH, and NINDS. The data used for the analyses described in this manuscript were obtained from dbGaP accession number phs000424.v8.p2 on 09/25/2023.

This research was made possible through the NIH Medical Research Scholars Program, a public-private partnership supported jointly by the NIH and contributions to the Foundation for the NIH from the Doris Duke Charitable Foundation, Genentech, the American Association for Dental Research, the Colgate-Palmolive Company, and other private donors.

## Web Resources

http://www.omim.org/

https://gnomad.broadinstitute.org/

https://www.ncbi.nlm.nih.gov/clinvar/

https://www.ncbi.nlm.nih.gov/snp/

https://muscleviz.github.io (Wittenbach et al., 2019)

## Data and Code Availability

**Supplemental Figure 1:** A) Reads aligned to Exon 22 in DNA show heterozygous SNP c.2079C>A; p.(Cys693Ter) present in 50% of reads. B) The variant c.2079C>A; p.(Cys693Ter) is present only in 11 out 64 reads in in RNAseq of P1,consistent with nonsense mediated decay prediction for this truncating variant

**Supplemental Figure 2:** analysis of all isoforms (no junction reads removed) existing in patient and controls between exons 142-145. The only difference between patient and controls is the lack of exon 144 inclusion. Note the 6 splice junction reads in patient starting in intron 144 and ending at exon 145. Patient in purple and three biceps controls in green, all adult (30-50yrs) biceps have a higher read count of exon 144 compared with 143 in adult biceps

**Supplemental Figure 3:** Patient compared with two age and sex matched controls, but not biopsied from the same muscle group. Both exons 143 and 144 are expressed almost equally in control samples, while the patient sample expresses exon 144 only. Again, Note the 6 splice junction reads in patient starting in intron 144 and ending at exon 145, which are not seen in controls.

**Supplemental Figure 4:** five randomly selected GTEX muscle samples biopsied from “below the patella” or “gastrocnemius” all show multiple rare isoforms, including isoforms splicing exon 142 directly to exon 145. Notably, there is no isoform that matches the six reads demonstrating intron 144 extension in our patient. There is variability in exon 143 expression between individual samples, although the lack of precise biopsy knowledge makes drawing conclusions regarding the relative abundance of 143 and 144 from GTEX data difficult.

