## Supplemental Tables 1-2, Supplemental Figures 1-4 for "Differential inclusion of *NEB* exons 143 and 144 provides insight into *NEB*-related myopathy variant interpretation and disease manifestation"

**Supplemental Table 1: Pathogenic variants affecting exon 144 in the literature.**

| Mutation | Second Hit | Clinical phenotype | confirmed Nemaline rods? | Citation |
|---|---|---|---|---|
| Pathogenic variants affecting exon 144 (NM_001271208) | | | | |
| ex144: c.21506C>A; p.Ser7169* | Homozygous | Unusual pattern of selective weakness, dystrophic biopsy. Difficulties in waking from anesthesia | YES | Lehtokari et al 2015 |
| ex144; c.21423del; p.Lys7141fs | int112; c.17737-2A>T; p.? | "Unspecified form of NM" | NO | Lehtokari et al 2015 |
| int144: c.21522+3A > G | no second hit identified | Presentation at 9yo with toe walking, no cardiac abnormalities, no other data available for this patient | YES | Lee et al 2017 |
| int144: c.21522+3A > G | Exon 81; c.12148G>T; p.E4050Ter | Childhood onset nemaline myopathy | YES | Wan et al 2019 |
| int144: c.21522+119C>G | Homozygous | Presented at 15 months with delayed walking | YES | Laflamme et al 2021 |

| | | | | |
|---|---|---|---|---|
| | | followed by proximal > distal muscle weakness. Facial/Bulbar weakness noted as well | | |
| Pathogenic variants affecting exon 144 but called exon 143 by NM_001164508.1 | | | | |
| in143:c.21417+3 A>G | ex 133 c.20360_20361insA; p.Thr6787fs | "typical congenital" | YES | wang et al 2020 |
| in143:c.21417+3 A>G | ex120; c.21793C>T; p.Arg5552* | "typical congenital" | YES | wang et al 2020 |
| in143:c.21417+3 A>G | ex45; c.5574_5575ins;p.Lys1859_Lys1860delins | "typical congenital" | YES | wang et al 2020 |
| in143:c.21417+3 A>G | ex18; c.1623delT; p.Asp542Ilefs*15 | "typical congenital" | YES | wang et al 2020 |
| in143:c.21417+3 A>G | ex124; c.19211delT; P.Leu6404Argfs*9 | "childhood onset" | YES | wang et al 2020 |
| in143:c.21417+3 A>G | ex109; c.17367G>A; p.Trp5789* | "childhood onset" | YES | wang et al 2020 |
| in143:c.21417+3 A>G | In80; c.12019-10G>A; | "typical congenital" | YES | wang et al 2020 |
| in143:c.21417+3 A>G | ex121; c.18917G>A; p.Trp6306* | "childhood onset" | YES | wang et al 2020 |
| in143:c.21417+3 A>G | ex61; c.8479C>T; p.Gln2827* | "childhood onset" | YES | wang et al 2020 |
| in143:c.21417+3 A>G | ex15; c.1263dupA; p.Tyr422fs | "Adult onset" | YES | wang et al 2020 |
| in143:c.21417+3 A>G | ex105; c.16465A>G; p.Lys5489Glu | "Adult onset" | YES | wang et al 2020 |
| in143:c.21417+3 A>G | ex53; c.7212T>G; p.Tyr2404* | "Adult onset" | YES | wang et al 2020 |
| in143:c.21417+3 A>G | ex115; c.18187C>T; p.Arg6063* | "typical congenital" | YES | wang et al 2020 |
| in143:c.21417+3 A>G | ex49; c.6195dupG; p.Tyr2066Valfs*4 | "childhood onset" | YES | wang et al 2020 |

| | | | | |
|---|---|---|---|---|
| in143:c.21417+3 A>G | ex61; c.8394T>G; p.Tyr2798* | "childhood onset" | YES | wang et al 2020 |
| in143:c.21417+3 A>G | ex3; c.36G>T; p.Glu12Asp | "childhood onset" | YES | wang et al 2020 |
| in143:c.21417+3 A>G | ex150; c.22037A>T; p.Asn7381Ile | "childhood onset" | YES | wang et al 2020 |
| in143:c.21417+3 A>G | ex128; c.19751T>G; p.Met6584Arg | "typical congenital" | YES | wang et al 2020 |

**Supplemental Table 2: Literature on distribution of exon143/144 mRNA**

| **Muscle** | **Donner et al 2006 MOUSE (143:144) qPCR** | **Donner et al 2004 HUMAN semi quantitative PCR** | **Laitila et al 2012 HUMAN Microarray and RT-PCR** | **Lam et al 2018 HUMAN qPCR** | **Uapinyoying et al 2020 MOUSE Long read sequencing** |
|---|---|---|---|---|---|
| **Gastrocnemius Medial** | D0: 1:0 ;D19 4:1; D21 1:1; W6 1:5 | Adult 144 only | | | |
| **Gastrocnemius Lateral** | D0: 1:0 ;D19 4:1; D21 1:1; W6 1:5 | Adult 144 only | | | |
| **Tibialis anterior** | D1 1:0; D21 1:1; W6 0:1 | Adult expressed both | statistically higher 143 than FDL, TP, FHL | | |
| **Extensor digitorum longus** | D0 2:1; D19 1:1; W6 0:1 | | Both - no relative expression data | | 100% 144 |
| **Soleus** | D4 1:1; D19 7:1; D23 4:1; W6 6:5 | | | | 63% exon 144; 37% exon 143 |
| **Vastus lateralis** | D23 1:1; W4 1:2; W6 1:14 | | weak exon 143 expression, good exon 144 | fast fibers: 143; slow fibers: 144 and 143 | |
| **vastus intermedius** | D23 1:1; W4 1:2; W6 1:14 | | | fast fibers: 143; slow fibers: 144 and 143 | |
| **vastus medialis** | D23 1:1; W4 1:2; W6 1:14 | | | fast fibers: 143; | |

| | | | | |
|---|---|---|---|---|
| | | | | slow fibers: 144 and 143 |
| **Rectus femoris** | D23 1:1; W4 1:2; W6 1:14 | Adult 144 only | Both are expressed, less 144 than adductor longus | fast fibers: 143; slow fibers: 144 and 143 |
| **Tibialis Posterior** | | | Less 143 than in the tibialis anterior, expresses both exons | |
| **Sartorius** | | | Both expressed, no relative expression data | |
| **Gracilis** | | | statistically lower 144 than adductor longus, expresses both | |
| **Adductor longus** | | | statistically higher 144 than gracilis, semimembranosus, biceps femoris, rectus femoris, and semitendinosus | |
| **Adductor magnus** | | | Both - no relative expression data | |
| **Semitendinosus** | | | statistically higher 143 than VL (both transcripts expressed) | |
| **Semimembranosus** | | | Expresses both transcripts, less 144 than AL but unclear if more 144 than 143 | |
| **Biceps femoris** | | | statistically higher 143 than VL, FDL | |
| **Masseter** | D0 1:1; D14 0:1; W6 0:1 | | | |
| **Diaphragm** | D0 1:2; D7 1:7; D21/W6 0:1 | | | |
| **Cardiac** | developmentally expressed equally | Adult 144 only | | |
| **Longus capitis** | exclusively 143 | | | |

### Supplemental Figure 1

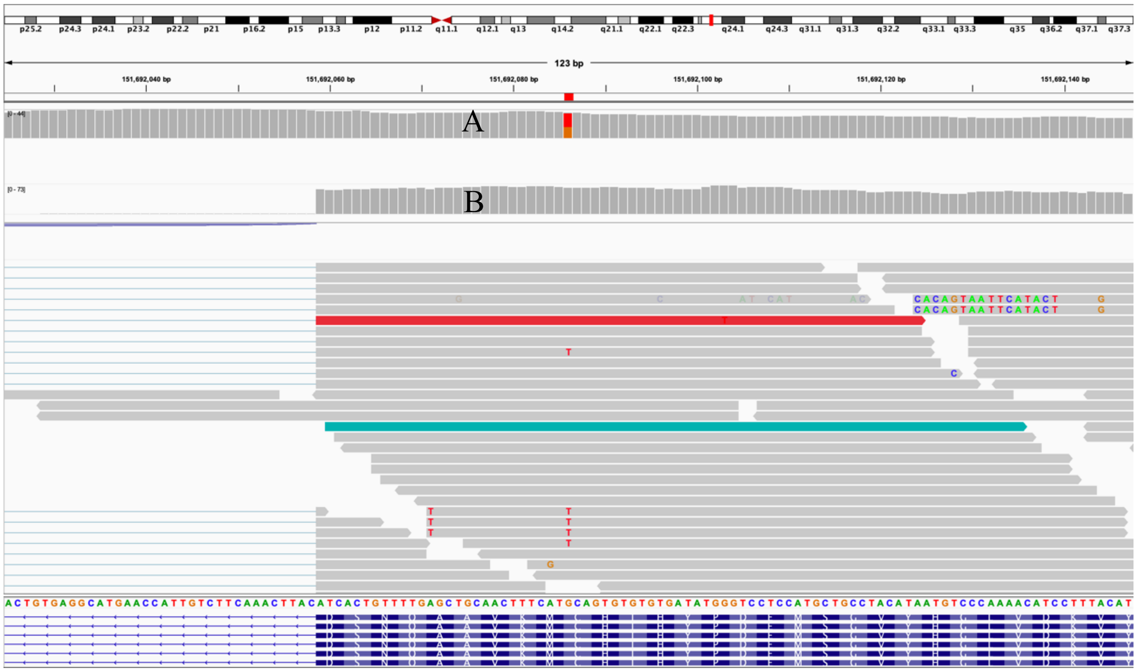

### Supplemental Figure 2

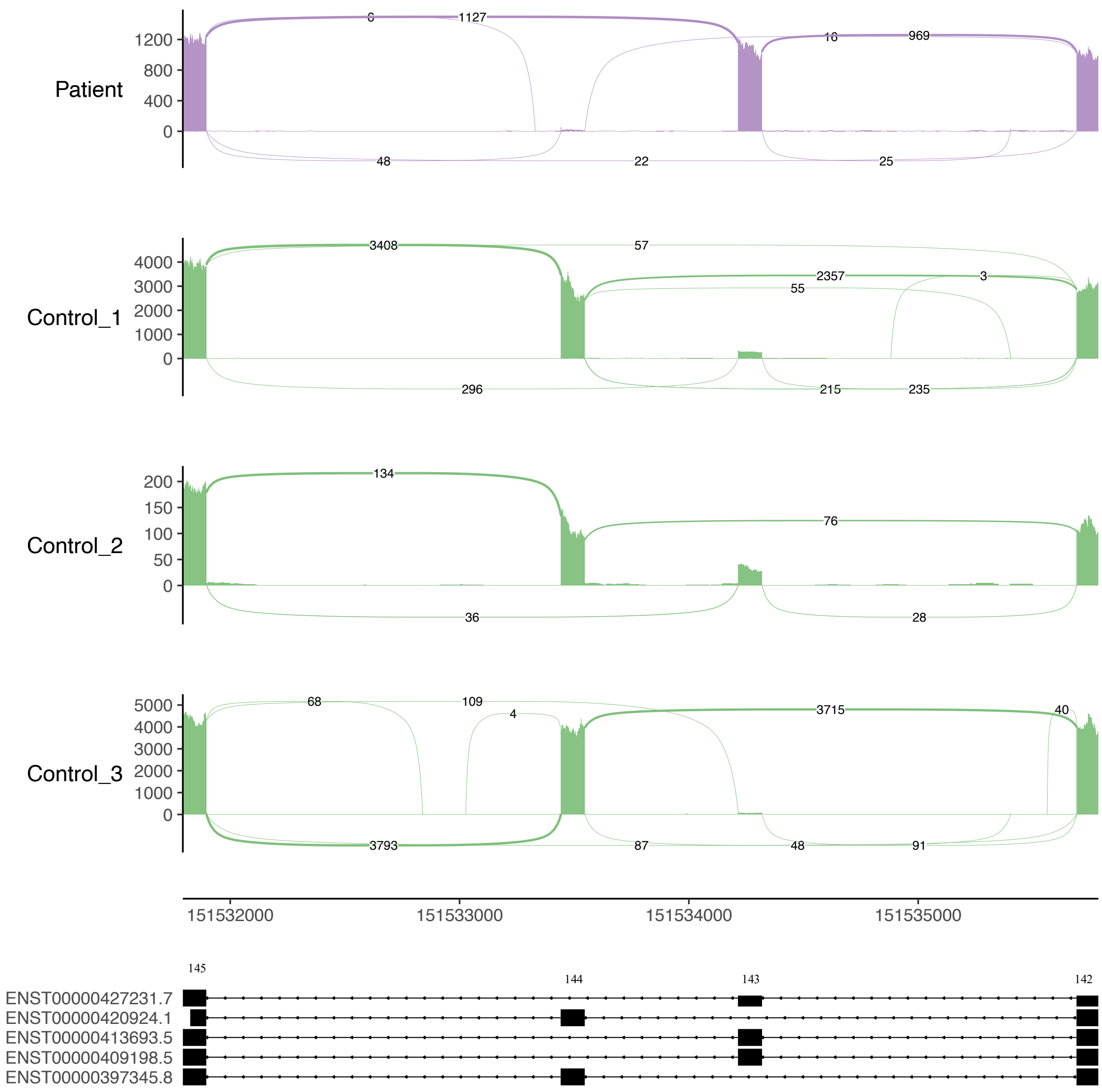

### Supplemental Figure 3

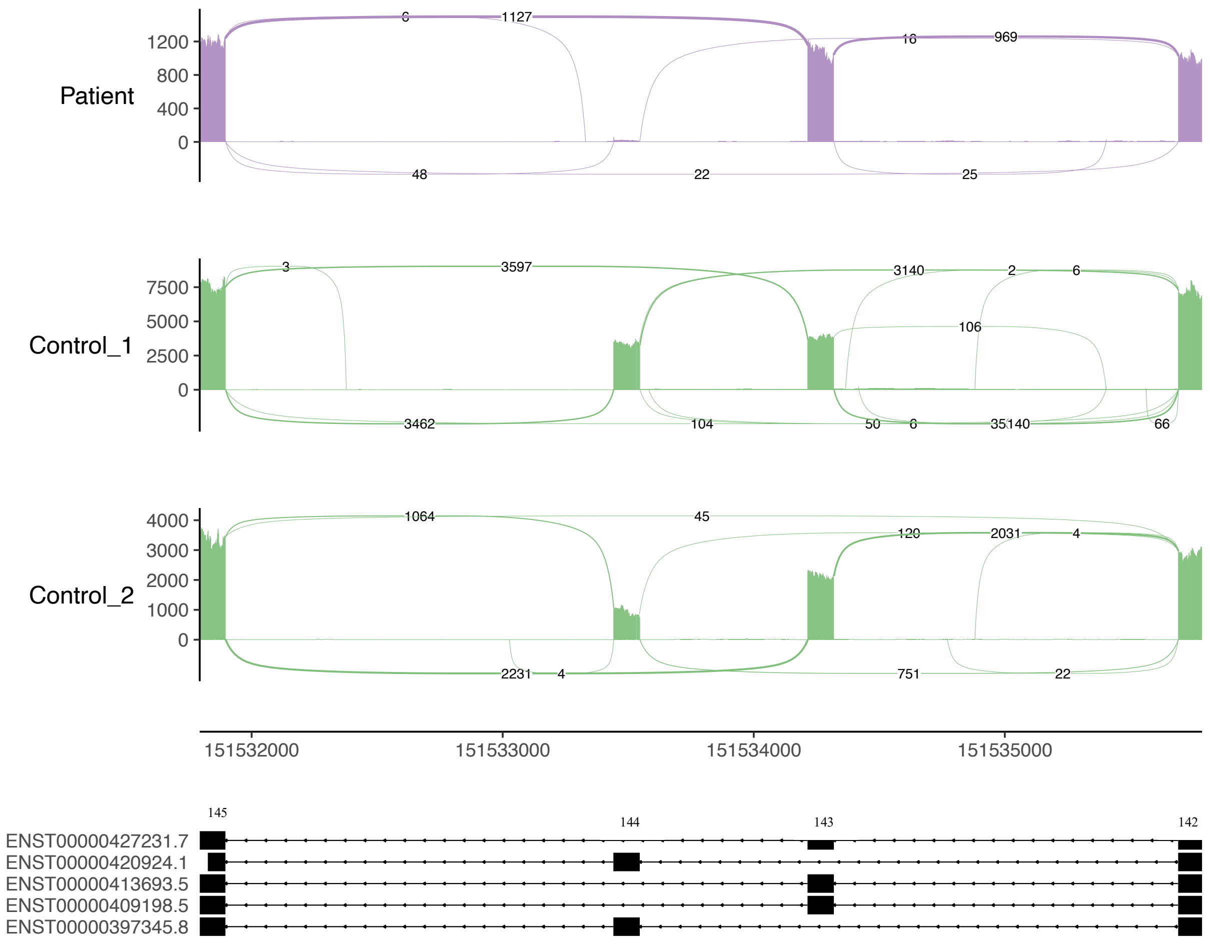

### Supplemental Figure 4

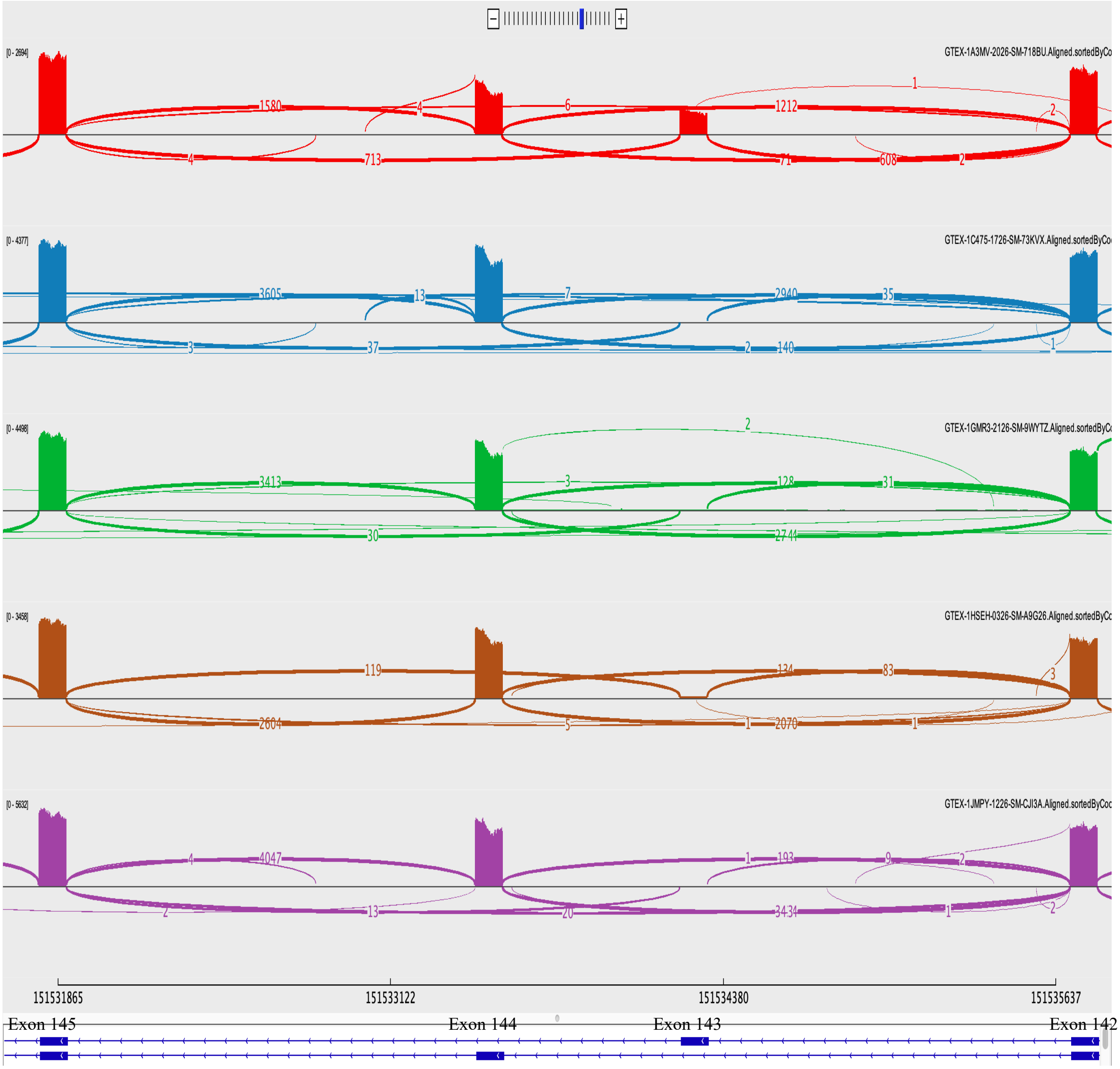
